# Climate change, livelihoods, gender and violence in Rukiga, Uganda: intersections and pathways

**DOI:** 10.64898/2025.12.01.25338207

**Authors:** Richard Muhumuza, Manuela Colombini, Pandora Zilstorf, Isla Collee, Gift Namanya, Joseph Katongole, Susannah Mayhew

**Author notes:** **Corresponding author:** Richard Muhumuza, MRC/UVRI & LSHTM Uganda Research Unit.

## Abstract

Climate change disproportionately affects poorer countries like Uganda, intensifying poverty and livelihood stress, which can escalate gender-based violence (GBV). This study, though not originally intended to focus on GBV, examines how it interconnects with poverty, shifting gender roles, alcoholism, environmental stress, and family planning dynamics.

Between April and July 2021, we conducted 28 focus group discussions (FGDs), comprising 6-8 participants each, stratified by sex and age (18–25, 25–49, and mixed 50+ groups). Additionally, 40 key informant interviews (KIIs) were held in Rukiga district, Uganda. Purposive sampling was applied. Data were organised in NVivo 12 and analysed thematically.

Participants described GBV, including intimate partner violence, non-partner sexual violence, child abuse, and early marriage, as widespread and normalised. Two main interconnected driver clusters emerged. First, poverty, male alcohol use, and shifting gender norms contributed to household instability. As men abandoned provider roles, women assumed more responsibilities, provoking conflict and sometimes violence from disempowered male partners. Second, environmental degradation and climate-related stressors (droughts, floods, soil erosion) worsened economic hardship, tensions, and GBV. Population growth and limited land access further strained livelihoods. While family planning was generally supported, male opposition sometimes triggered conflict.

Climate change impacts are gendered, with GBV pathways shaped by shifting gender roles, norms, and relations destabilised by environmental and livelihood pressures. Addressing GBV in climate-affected communities requires gender-transformative environmental and livelihood programmes. This should include strengthened social and structural resilience to challenge inequitable gender norms and power imbalances.

## Introduction

Climate change is a global phenomenon, though its impacts are not equitably distributed (1) with the poorest countries of the world, including Uganda, experiencing the most adverse effects (2, 3). The effects of climate change within Uganda are evident and are projected to increase exponentially (4), causing changing weather patterns and increased occurrence and severity of natural disasters, such as droughts, floods and landslides (5–7). These events lead to widespread land degradation and damage to infrastructure, settlements and the agricultural sector, causing substantial economic and livelihood loss (8). Uganda’s vulnerability to climate change and climate-sensitive disasters is extremely high and is influenced and exacerbated by extreme structural, political and economic fragility and a high dependence on climate-sensitive sectors, including the agricultural sector (5).

The effects of climate change and environmental degradation, and the subsequent vulnerability of people, cannot be separated from the indirect and socially mediated effects, which intersect with wider social, cultural and economic factors (9, 10). Indirect effects include the exacerbation of poverty and livelihood stresses, which in turn can escalate gender-based violence (GBV) (11, 12). Gender is among the most universal and important factors of vulnerability to the effects of climate change, as demonstrated in numerous systematic reviews (12–15). Owing to inequities in the distribution of power and access to social and material assets, such as food, water, land, education, health, housing, employment and social capital, women and girls are more vulnerable to ecological and financial disruptions caused by climate variability (16, 17). Climate change stressors exacerbate pre-existing gender inequalities that expose women to greater risk of poverty, marginalisation and GBV (15). GBV is a major public health and human rights issue globally, affecting one in 3 women (18). In Uganda, between 45-56% women aged 15-49 have experienced physical and/or sexual violence (Uganda DHS, 2018). The most vulnerable women are those in rural areas, those with lower levels of educational attainment, and women from low-income households (Uganda Bureau of Statistics/Ministry of Gender, 2019). Efforts to reduce levels of GBV are evident in national policies, such as the 2016 National Policy on Elimination of Gender Based Violence in Uganda, and commitment to the 2021-2025 National Action Plan for Women, Peace and Security. However, action in the policy arena is undermined by high levels of social acceptability of GBV. According to the 2016 Uganda Demographic Health Survey (DHS), almost half (49%) of women aged 15-49 agree that a husband is justified in beating his wife for reasons such as burning food, refusing sexual intercourse, or failing to care for children (Uganda DHS, 2018). GBV leads to multiple short and long-term adverse consequences for women and their children. In particular, GBV can affect physical and mental health and social well-being (19–22). GBV is also a major contributor to reproductive health issues, including unwanted pregnancies and sexually transmitted infections (23, 24).

The intersectionality of climate change with gender roles, empowerment, and GBV presents complex challenges for development practitioners and policymakers (19, 25, 26). The existing literature reveals the links between climate change, environmental degradation, and GBV to be complex and highly context dependent. Previous studies have demonstrated how drought increases the risk for GBV throughout SSA, how food insecurity is associated with intimate partner violence in central and western Uganda, and similarly, how disasters triggered by climate change are associated with GBV (27, 28). Several factors are thought to play a key role in mediating these relationships, including poverty, food insecurity, water scarcity, loss of livelihoods, male unemployment, and alcohol abuse (14, 29). Alcohol use (often triggered by poverty-related stress) is known to be a consistent risk factor for GBV, and harmful use increases the risk of violence and injury to women and children (30). Environmental stressors may also exacerbate pre-existing risk factors for GBV, such as socioeconomic inequality, gender discrimination, rigid sociocultural gender norms, and power imbalances at various societal levels (14). Gender norms, roles, identities and relations are increasingly being recognised as important mediators between environmental stress and violence, as they are often destabilised during times of crisis (31). For example, threatened livelihoods and increased financial and food insecurity may increase the inability of men and boys to uphold the socially ascribed role of ‘provider’, increasing their likelihood of perpetrating violence against women and girls (32–35).

While the nexus between gender inequality and climate change is increasingly acknowledged and becoming a focus of international policy and practice frameworks, and research (17)Most attention has been on the impact of climate disasters and natural hazards, such as floods, droughts, and earthquakes (14). The impact of chronic slow-onset climate change, like that being experienced in Rukiga, is neglected, and the indirect pathways by which climate-related factors lead to GBV in these settings remain understudied (36).

The purpose of this study is, therefore, to examine locally held perceptions of the relationship between climate- and livelihood-related stressors and changing dynamics, including the risks of GBV in the Rukiga District of Southwestern Uganda.

## Materials and Methods

### Study setting

Rukiga District in western Uganda experiences slow-onset, chronic climate and environmental change. It features steep upland slopes and low-lying wetland areas, which serve as the habitat for Uganda’s national bird, the Endangered Grey Crowned Crane. Most residents in Rukiga District engage in small-scale farming, traditionally on the steep valley sides and increasingly in wetland valley bottoms. Rising human activity is exerting greater pressure on local ecosystems, resulting in degraded wetlands and declining soil quality in upland farms. Additionally, access to health and especially family planning services remains limited despite high demand for these services. This was a formative study using qualitative research methods in four parish communities within Rukiga District (Nyakagabagaba, Kitojo, Kihanga-Sindi, and Burime).

### Data collection

From 20 April 2021- 02 July 2021 five focus group discussions (FGDs) were conducted in each community (28 in total) each consisting of four to six participants, stratified by both sex (male and female) and age (18-25, 25-49, and 50+ years), except for the one of 50+ years which was mixed with an equal number of males and females. After the FGDs, 40 Key Informant Interviews (KIIs) (10 per community) were conducted. Participants were purposively selected (excluding those who had participated in the FGDs) to represent different age groups and sexes from the community with the help of community leaders. Written informed consent was obtained from the participants before participation in the study. The FGDs and KIIs were conducted within the study villages in an environment that was private for both the participants and the researcher, including schools and church buildings. Experienced qualitative researchers (one male, RM, and one female GN) conducted the FGDs and KIIs, each lasting between 60-120 minutes. A semi-structured guide was used to gather information on the problems affecting the community and the possible connections between these problems. It did *not* include specific questions or probes on GBV since the project was not originally focused on this.

### Data analysis

FGDs and KIIs were conducted in Runyankore/Rukiga, audio-recorded, transcribed verbatim, and translated into English. The transcripts were cross-checked by the research team against the audio recordings to ensure accuracy. GBV was not the focus of the study, which is described elsewhere (34, 37), but because mention of it was ubiquitous across the sites, we undertook a specific analysis of what participants said about GBV. Originally, the FGDs were intended to capture community norms and responses to health and livelihood challenges. In contrast, the KIIs were intended to triangulate these community norms (from a “key informant” perspective) and to capture information on the nature of any existing collective and institutional responses to the challenges identified. In the initial identification of GBV as an emerging issue, it was clear that there was little difference in the way that FGD and KII participants spoke about GBV therefore for the GBV-specific analysis reported in this paper the two datasets were analysed together using the same code frame.

Transcripts were coded and organised using NVivo 12 and analysed thematically, drawing on the research questions as well as emergent themes (38, 39). The analysis was an iterative process of discussion and revision between the wider research team. Three researchers generated a list of recurrent codes by independently reviewing transcripts several times and making notes of key ideas and codes. After completing the initial round of coding, the researchers scrutinised the new codes and comparisons were made to check and ensure consistency. Discrepancies in the coding were re-examined, and an initial coding framework and codebook were developed and used after consensus on the final codes. Data were organised by assembling emergent themes and identifying recurring patterns and categories in context to the research questions. We present the overall findings from the FGDs and KIIs organised by the final themes.

### Ethical Considerations

We obtained informed written consent from all literate participants. For participants who were not literate, a witness confirmed that they had understood the purpose of the study. The witness was somebody who was chosen and trusted by the participant.

## Results

We first briefly describe how openly respondents spoke about GBV during the research. We then present respondents’ perceptions of the interconnected causes of and pathways to GBV. These emerged in two main clusters. First, environmental degradation and stressors interplay with livelihoods and family planning use (which also connect back to poverty). Second, a complex interplay between lack of income (poverty), changing gender roles (with women becoming breadwinners) and alcoholism.

### Ubiquitous awareness of Gender-based violence

As noted in the methods, the original project was not set up to research GBV, and the topic guides contained no prompts about it. Despite this, GBV was spontaneously mentioned in almost every FGD and KII, emphasising its pervasiveness. Forms of GBV mentioned included non-partner sexual violence such as rape, intimate partner violence (physical, sexual, and emotional), violence towards children, and early marriage and unintended pregnancies. One key informant mentioned femicide. Despite a small number of men describing women being abusive towards husbands or children, the majority of participants (both men and women) recognised that men were the primary perpetrators of physical harm towards women and children, respectively.

Throughout our data, GBV was widely acknowledged as a pervasive individual, family, and community issue that was normalised in people’s lives. Although GBV was widely regarded as a norm, it was also widely acknowledged as a “problem” that negatively affects people. This is underscored by the euphemisms for GBV that many participants used, such as “no peace”, “quarrels” or “conflict” throughout their experiences and perceptions.

GBV was raised during the discussion of a wide range of factors, which are now discussed in terms of the two main emerging clusters of interrelated pathways.

### Climate-related environmental and livelihood stressors

In every village, participants spoke of the devastating effects that environmental and climatic stressors were having on their livelihoods, health, and well-being. Challenges included drought, flooding, landslides, soil erosion, reduced soil fertility, diseases affecting crops and livestock, and unpredictable seasonal and rainfall patterns.

> *The climate normally disturbs this village. Floods normally give us a hard time. Whenever it rains, because of the hills, all the soil is eroded, and all the crops get destroyed. KII Male, 25-49 years Village B*

> *Climate change affects us. There are times when it shines a lot. This causes problems in the season because crops will be destroyed by a lot of sunshine. Other times, you find that when it rains heavily, crops also get destroyed.” KII Female, 25-49 Years Village A*

Respondents spoke of how these climate-related challenges affected agricultural yields, which meant there was less food and less income. Respondents described how this disruption to their livelihoods led to family instabilities and conflict, demonstrating how GBV is triggered or exacerbated by the undermining of livelihoods that is caused by climate and environmental challenges like flooding and landslides:

> *“On top of famine, even the soils change. They lose fertility because the top fertile soils are washed away. […] When you cultivate and fail to yield well, you cannot have income. […] It causes us poverty and unrest in families because, in the absence of money, you cannot have peace in a home.” KII Female, 25-49 years Village A*

Many participants spoke of these connections. A participant in one FGD of younger women clearly articulated the connections she saw between the landslides, which destroyed her subsistence farm, leading to a lack of food, which caused her husband to beat her in anger. In her mind, the “chaos” was the direct result of her farmland being destroyed by landslides:

> *When the environment gets worse, say, if landslides destroy my garden, that means that even if I cultivate there, I will not get what to eat. Then, the moment I fail to get what to cook for the family, now, even if the man is away and comes back and finds that I have not cooked, he will say, my children are starving, anger makes him beat the woman, the woman transfer the anger to children and also beat them, now you find the whole family chaotic not because you lacked what to cook but because the land was destroyed by landslides. FGD Females, 18-25 Years Village C*

Another often-mentioned factor that affected the loss of productive farmland and put further pressure on livelihoods was the ever-growing population, meaning that large families can no longer produce sufficient food to feed themselves.

> *People in ancient days […] had plenty of land and would cultivate, let their soils rest, and regain fertility, but now, maybe because of the overproduction of children, the land is scarce. So, I see this as a great problem, and it is getting worse.” FGD Males, 25-49 years Village A*

This pressure further exacerbated the livelihood stresses caused by climate change in two different ways: by multiplying the number of people dependent on the same tract of land, and by leading to environmentally unsustainable farming practices.

> *On the environment, people cut down all the trees on the hills, leaving the land bare. When it rains, water from the hills combines with that from the houses, and it floods the valleys, destroying the crops. When a person has nothing to eat, that’s when they go and burn the wetland to catch mudfish for food. KII-Male-25-49 Years-Village D Even if a person tries to dig and grow crops, due to many children to feed, he cannot get any food balance to take to the market for sale. They eat it all. Because they do not have an alternative, they end up encroaching on some reserved lands and cultivating them. KII-Male-25-49 Years-Village B*

> *Because of limited land and growing crops on the same land, the soil loses fertility. So even when you plant your crops, they don’t yield, and when the sunshine comes, they all dry up and decay in the rainy season … What causes that is poor farming, wetlands were seriously cultivated, and when there is water runoff, it affects everywhere. People don’t mind planting trees that reduce the amount of water. FGD-Females-25-49 Years-Village A*

One long-term solution that many respondents were keen to have is better access to family planning (FP):

> *“[…] you should make sure you get us family planning methods that fit our health so that our families are put in good condition.” FGD Females 25-49 years Village B*

Yet disagreements surrounding women’s use of family planning methods were raised by many participants as a specific source of conflict between women and their husbands, although most respondents (men and women) expressed positive attitudes towards FP. Where men did not support FP, the reasons for their rejection included a desire to have more children and a lack of understanding of FP methods. One woman explained that:

> *“People understand differently. Some like and support it [fp] while others do not support it. Still, some women do family planning secretly without the knowledge of their husbands.”* FGD M/F, 50+ Years Village D

Whilst some women overcame opposition from their husbands by accessing FP services in secret, the *possibility* of conflicts over FP usage acted as a deterrent to other women.

> *Most women do not tell their husbands about family planning. They just wake at once and go for family planning without telling their husbands… as the man discovers it, they both resort to quarrels. Now, as their colleagues discover this, they also fear joining family planning* FGD M/F, 50+ Years Village D

### Poverty, alcohol use, and changing gender roles

When describing their experiences and perceptions of poverty and its associated consequences including poor diets, sickness, and lack of ability to pay for healthcare and transport to medical facilities, most respondents explicitly identified poverty as a direct cause of GBV:

> *“…when poverty knocks on your door, you start quarrelling and fighting and consequently, the family ends like that”.* KII Female, 50+ Years Village C

> *When a child gets sick, you do not have money. When you fail to get food, you fight with the woman. So, I think failure to get daily income is the leading cause of our problems.” FGD Males, 25-49 Years Village B*

Poverty was also seen as a trigger for alcohol use: “*You find that the man has no income. He has resorted to spending all his time at the bar”* (FGD Female, 18-25 years Village C). Harmful alcohol consumption was in turn, described as a direct trigger for GBV:

> *“When he comes home drunk and he talks, then you keep silent, he beats you …. So, alcoholism also causes problems.” KII Female, 18-25 years Village B*

> *“And men who drink alcohol, in their homes, there is a lot of quarrels.” KII Female, 25-49 years Village A*

A common narrative emerging amongst participants was that male harmful alcohol use directly increased household financial and economic instability. Through spending the family’s already limited income on alcohol, men redirected income away from food, basic household goods, and children’s school fees. Some respondents described how men steal their wives’ harvests or income to purchase alcohol. This was cited as a major source of conflict between spouses:

> *“The money that is intended to support the family is often found to have been spent at the bar, leading to conflicts within the family.” FGD Males, 18-25 years Village C*

Poverty was not only identified as a consequence of harmful alcohol consumption but also a cause, largely mediated through unemployment and feelings of idleness. In this way, respondents painted a picture of a vicious cycle between poverty, alcohol dependency, and GBV:

> *Men don’t have [anything] to do, so for every coin they get, they direct it to the bar. If they had jobs and they got the money, they would be planning for their families. So, poverty is one cause of alcoholism. KII Male,* 50+ years Village D

> *“Development in the homes of men who drink a lot of alcohol is very low […] those who stay in the bar, cannot do anything in their homes” KII Female, 25-49 years Village A*

Alcohol use and GBV were also discussed in relation to changing gender roles and norms. From the outset, participants gave clear accounts of very traditional and patriarchal roles of men and women, where men were seen as the main, respected decision-makers. By contrast, the roles and expectations for women see them as homemakers and responsible for the well-being of the children and family.

> *“The men are the ones who are supposed to make decisions because they have more respect than the women”* FGD Female, 18-25 years Village D

> *“We know men are heads of families, but we women have a lot of activities in the home.” FGD Females, 25-49 years Village B*

Yet, despite these traditional characterisations of gender roles and expectations, respondents described a changing reality in which men were accused (by both men and women) of abandoning their duties as family heads with responsibility for income and accompanying decision-making shifting to the shoulders of the women:

> *Under normal circumstances, the head of the family is the man. But these days, you find a man does not want to know about the well-being of his family. The woman wakes up early and prepares for the children to go to school, and the man does not want to know. It is the woman that takes care of everything.” FGD Females, 18-25 years Village A*

To provide an income able to support their families in the absence of male breadwinning, many participants described the extra efforts of women to provide for their families, often characterising alcoholism as a cause of women rising to take up abandoned responsibilities.

> *“A man wakes up early and goes to the bar, fails to go to cultivate [the farm] with his wife, and drinks the whole day. Isn’t [it] here that senses disappear? So, that is why a woman leads such a family because such a man is senseless.” FGD Males 50+ years Village B*

> *“Even in our families, change came because when a woman finds out that her husband is an alcoholic and not responsible, she takes up the responsibility of the family, digs for money, and pays school fees for her children, can’t wait for the man to do it. So, change came and now women woke up.” KII Female, 25-49 years Village A*

Meanwhile, the increasing engagement of women in income generation for their families threatens the traditional power balance, with participants describing the male partners of such women as lacking authority.

> *At times, you find that you have a wife who has a business and has more money than you. So, such a woman will rule you because, for you, you stay in the village and have nothing to do.” FGD Males, 18-25 years Village B*

> …*the man is only thinking about alcohol, when the child asks for a book, the father tells the child to ask the mother because the mother is the one who works, and therefore, she has some money. So, the man does not have the authority to issue some directives in his home just because he does not have money.* FGD Female, 18-25 Years Village A.

However, women stepping into the role of breadwinner does not earn her the respect that is traditionally given to men in such positions. Rather, she is regarded with suspicion because the switching of roles is uncomfortable, undermining the position of men as the quotes above indicate in their phrases “such a woman will rule you” and “the man does not have the authority to issue some directives in his home”. This suspicion was reported to result in serious, violent consequences for some women, as though men could not accept that women could legitimately earn money through hard work and efficiency but must instead have been unfaithful – the ultimate humiliation for a husband:

> *“You find that the man has no income. He has resorted to spending all his time at the bar. Now, for me, when I cultivate a given garden, I keep saving some money so that when it accumulates, I can also buy some land for cultivation. When the man discovers that you have bought a piece of land, he can nearly beat you to death. He asks you where you got the money from. Did you adulterate for it? Now, you find instabilities rising.” FGD Female, 18-25 years Village C*

Yet the impact of such violence would not only be felt by the women, but the whole family, as this male participant explained:

> *It does not only disturb the woman, but also the income of a home dies completely when domestic violence happens. KII* Male, 25-49 Years Village C).

These extracts indicate how alcoholism can be fuelled by poverty, triggering GBV, but also how alcohol abuse is connected with changing gender roles. Alcoholism was seen as both as a cause and a consequence of perceived male disempowerment and a failure to meet traditional markers of masculinity. Consequently, this led to women taking leading roles as decision-makers and breadwinners for their families, but often with violent consequences.

## Discussion

This study examined locally held perceptions of the relationship between climate- and livelihood-related stressors and changing gender dynamics, including the risks of GBV in the Rukiga District of Southwestern Uganda. Our findings reveal a complex interplay of factors, including climate-related livelihood stressors, poverty, harmful alcohol use, change in gender roles and norms and gender-based violence. The perceptions of the interconnections, as reported by our respondents, are summarised in Figure 1.

**Figure 1:**
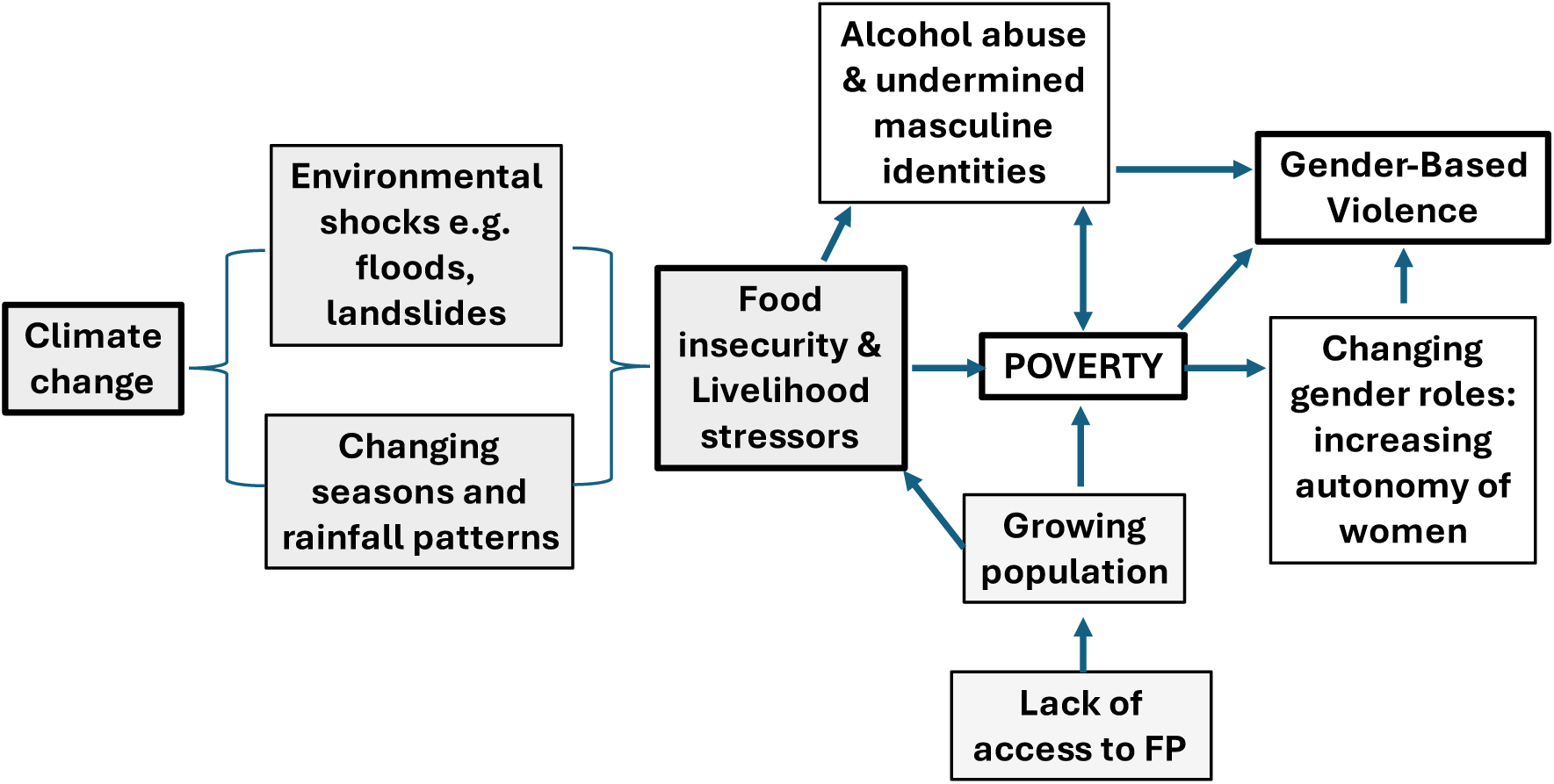
Perceived connections and pathways between climate change, livelihoods and gender

Poverty was seen as a pervasive direct trigger for GBV, and climate change exacerbates poverty through environmental shocks and chronic change that leads to food insecurity and livelihood stressors. Growing populations, partly because of a lack of access to FP services, also contribute to food shortage and poverty. Poverty was seen as a trigger for harmful alcohol use, which was also identified as a direct cause of GBV, and particularly of partner violence against their wives. In the context of rising poverty and increased male alcohol abuse, many women were taking on income generation roles themselves in order to feed, clothe and educate their families. These changing gender roles in relation to women’s economic independence were seen to result in a concomitant reduction of male decision-making roles. This also led to family tensions that were sometimes described as resulting in GBV. We discuss these interlinkages in relation to wider literature.

### Interlinkages between climate change, environmental and livelihood stressors, and poverty

The findings of our research demonstrate how climate change, extreme environmental fragility and economic and livelihood insecurity are inextricably linked. When combined, they exacerbate one another to increase poverty. Poverty emerged as the most prominent direct trigger for GBV by creating acute and chronic household and community stress, thereby indirectly linking climate change to GBV. These findings support the work of others, notably Carney et al. (2020), who show how the effects of both chronic climate change (prolonged dry seasons) and climate induced environmental disasters in Uganda led to the destruction of natural resources that underpin livelihoods, driving resource scarcity and poverty, thereby leading to increased risk of GBV (34, 40). A systematic review of natural hazards and disasters and their link with violence against women and girls (14, 37) identified important pathways which, though not specific to chronic or slow-onset climate change, do mirror our findings. In particular, they find that extreme climate events increase economic stressors that lead to violence and exacerbate existing drivers of GBV. Other work in Uganda, not specific to climate, also identified poverty and economic insecurity (41) as well as alcohol abuse as significant drivers of GBV (42). Livelihood pressures and alcohol abuse are well-known triggers of violence, evidenced in wider literature across multiple settings, including sub-Saharan Africa (35, 43).

### Alcohol abuse and masculine identities

Many respondents identified men’s alcohol use as a key factor mediating the relationship between environmental degradation, livelihoods and violence. This is supported by Uganda’s most recent DHS data, which found that women whose husbands are often drunk are much more likely (84%) to experience GBV than women whose partners do not drink (45%) (44). Whilst a consensus exists in the literature that alcohol itself is not a “necessary or sufficient cause” of GBV (45, 46), it is recognised that alcohol use, especially excessive consumption, influences the frequency and severity of GBV (47, 48). However, the mechanisms underlying this association are still contested. In this study, participants recognised alcohol abuse by men to be both a symptom and cause of financial stress, unemployment, and feelings of inadequacy following a failure to fulfil their gendered role of provider. Adopting a gendered perspective illuminates how alcohol misuse may intersect with gender norms to elicit violence, such as in contexts where drinking is associated with harmful notions of masculinity (46), or used as a coping mechanism. In these contexts, alcohol use may be used to assert harmful masculine identities, especially in the absence of other traditional denotations of masculinity, such as employment (46).

### Changing gender roles: women’s economic participation

The participants in our study relied heavily on agricultural practices and natural resources for their livelihoods, and the disruptive impact of climate change on these sectors led to destructive social and economic systems. Participants illustrated how environmental stressors, including crop failure and loss of livelihoods, were forcing women to adopt new roles and responsibilities out of necessity. As men turned to alcohol in frustration, women increasingly acted as primary income earners, with increased decision-making power over household resources. Participants also described how these gender role reversals were a source of household conflict and led to partner violence. This is consistent with other literature, which shows that when the social fabric and corresponding gender roles within communities are dramatically altered by the deterioration of local economies, dispossession of land, loss of local livelihoods and degradation of natural resources, this can give rise to GBV (Alston & Whittenbury, 2012; Weltbank, 2012; Namasaka, 2014; Barcia, 2017b). Carney et al. (2020) highlight how livelihoods disruption can cause social vulnerability, community wide livelihood and economic insecurity and acute and chronic stress, which strengthens the conditions for gender-based violence to occur (40). Previous research also suggests that GBV is common in transitional societies undergoing sociocultural shifts away from traditional patriarchal values towards gender equality. This has been studied in the Ugandan context, where domestic violence in Wakiso District was found, in part, to be a response to unequal balances of power in a shifting socio-economic environment (49) and appears to be what we are seeing in Rukiga as well.

Our findings contradict other literature, however, that suggests natural disasters may induce positive shifts in gender relations through enabling increased female economic participation in post-disaster settings (50), which some authors have seen as acting as potential ‘windows of opportunity’ to drive social change (31, 51). Whilst women’s economic empowerment interventions, such as microfinance programs and cash transfers, have been suggested to lower women’s GBV risk (52, 53), evidence for their effectiveness is varied. Evidence from the MAISHA study for prevention of intimate partner violence (IPV) in Tanzania found that whilst an overall increase in women’s income was protective against IPV, those who financially contributed more than their husbands were at greater risk of sexual and physical violence (54). These effects may be explained with reference to gender theory. *Gender role strain theory* hypothesises that men who perceive themselves as failing to fulfil expected male duties, such as providing for their families, experience poor psychological outcomes, which may increase infliction of violence against their partners (55). Similarly, *gendered resource theory* posits that in traditional households where women act as breadwinners, husbands use violence to compensate for feelings of disempowerment (56). Both may be applied here to explain how increased economic participation by women in Rukiga challenges hegemonic masculine identities.

Despite the challenges of GBV faced by women like those in Rukiga, our findings show that they possessed considerable capacity to adapt to environmental degradation and mitigate effects on livelihoods. Adaptive strategies outlined by participants included adopting the dual burden of caretaker and primary income earner, engaging in alternative livelihoods, and joining savings groups. These findings support a growing body of research that identifies women as key agents of adaptation to climate change-related challenges (31). Nevertheless, the invisible yet dangerous pathways to GBV that we identify must be carefully considered by programs seeking to empower women through encouraging economic participation, to avoid exacerbating pre-existing challenges.

### Limitations

Two main limitations must be acknowledged. Firstly, as GBV was not an initial focus of the parent study, this data provides an incomplete picture of GBV in Rukiga district. Further qualitative research specifically investigating GBV must be conducted to further understand the complex pathways between environmental degradation, climate change and violence.

Secondly, the effects of sampling, social desirability, and recall bias must be considered in all qualitative research. Here, key informant interviews (KIIs) were conducted with individuals in positions of power within their communities. Given that experiences of gender-based violence (GBV) differ according to social status, poverty, and education, these perspectives may not represent those of less-powerful individuals, although conducting focus group discussions (FGDs) also helped to mitigate this risk. However, potential imbalances in wealth and educational power may have been evident between the interviewers and participants, particularly in the FGDs, which could have facilitated acquiescence and social desirability bias, where participants felt compelled to respond in certain ways. Again, triangulation with the KIIs assisted in mitigating this, and we noted the similarities in responses. Nevertheless, further research is necessary to adequately understand the breadth of experiences of GBV in Rukiga.

## Conclusions

As the effects of climate change continue to threaten global human and environmental health, it is increasingly important to understand how environmental changes will disproportionately impact vulnerable populations and exacerbate pre-existing issues. Our findings revealed a strong perception of worsening household and livelihood stress linked to climate change, which seems to be reshaping gender roles and dynamics within the family and, in some cases, contributing to increased conflict within households.

This demonstrates the importance of considering the gendered impacts of climate change in vulnerable communities, building social and structural resilience to effectively prevent GBV. Promoting environmental and livelihood programmes that are gender-transformative would help to address gender norms and power inequities so that both men and women are able to support their families, despite the challenges of climate-change and environmental degradation.

## Declarations

## Data Availability

Data will not be shared publicly due to the data-sharing policy of the LSHTM requiring a prior data-sharing agreement. However, the dataset containing the data supporting the study findings in this report can be obtained upon request from the corresponding author.

## Acknowledgements

We acknowledge the contributions of the MRC/UVRI and LSHTM Uganda Research Unit, the Margaret Pyke Trust, Rugarama Hospital, the International Crane Foundation, and the Endangered Wildlife Trust for their support services, which made this study possible. We are particularly grateful to all the study participants, not forgetting the mobilisers, for the time and information they shared with us.

## Author contributions

**Conceptualization:** Richard Muhumuza.

**Data curation:** Richard Muhumuza, Gift Namanya.

**Formal analysis:** Richard Muhumuza, Gift Namanya, Joseph Katongole, Isla Collee, Pandora Zilstorf.

**Funding acquisition:** Susannah Mayhew.

**Methodology:** Richard Muhumuza, Gift Namanya, Susannah Mayhew.

**Project administration:** Richard Muhumuza.

**Resources:** Richard Muhumuza, Susannah Mayhew.

**Supervision:** Richard Muhumuza, Susannah Mayhew.

**Writing – original draft:** Richard Muhumuza, Gift Namanya, Isla Collee, Pandora Zilstorf, Joseph Katongole, Manuela Colombini, Susannah Mayhew.

**Writing – review & editing:** Richard Muhumuza, Gift Namanya, Isla Collee, Pandora Zilstorf, Joseph Katongole, Manuela Colombini, Susannah Mayhew.

